# Phase I Dose Ascending, Safety and Pharmacokinetics Study of APC148, a Novel Metallo β-Lactamase Inhibitor in Healthy Volunteers

**DOI:** 10.64898/2026.03.24.26349153

**Authors:** Bjørg Bolstad, Ragnar Hovland, Johan Bylund, Erik Rein-Hedin, Sandra Kuusk, Bjørn Klem, Pål Rongved

## Abstract

APC148 is a novel metallo-β-lactamase inhibitor with broad activity against Ambler class B enzymes including NDM, VIM and IMP. It is being developed for patients with serious infections caused by multidrug-resistant Gram-negative bacteria. APC148 is combined with the broad-spectrum β-lactam antibiotic meropenem and the serine-β-lactamase inhibitor avibactam, which targets Ambler class A, C, and some class D (OXA-48-like) enzymes. In combination with meropenem and avibactam, APC148 demonstrated superior *in vitro* activity against a global, multidrug resistant collection of Enterobacterales, showing its promising activity against β-lactamase producing pathogens.

In this randomized, placebo-controlled, first-in-human study, the safety, tolerability and pharmacokinetics of APC148 were evaluated in healthy adults. Single doses ranging from 50 mg to 760 mg APC148 were administered intravenously over 3 h to 46 participants across six dose groups. APC148 was well tolerated at all dose levels. All adverse events were of mild intensity, and no serious adverse events or adverse events leading to study- or treatment discontinuation occurred. The pharmacokinetics of APC148 were dose-proportional with low plasma clearance, low to moderate volume of distribution and a mean plasma half-life of 2.6 h.

APC148 is well tolerated in humans at therapeutically relevant doses and represents a promising candidate in the fight against antibiotic-resistant bacteria. (This study has been registered at ClinicalTrials.gov under registration number NCT06360640).

## INTRODUCTION

Infections caused by antimicrobial resistant pathogens represent a significant and growing global disease burden. Between 2025 and 2050, 39.1 million deaths attributable to antimicrobial resistance (AMR) and 169 million deaths associated with AMR are forecasted (1). Carbapenem-resistant Gram-negative Enterobacterales, including *K. pneumoniae* and *E. coli* are ranked among the highest-priority bacterial pathogens by the WHO (2).

Broad-spectrum β-lactam antibiotics such as carbapenems (e.g. meropenem) and cephalosporins (e.g. cefepime) are hydrolyzed by serine- and metallo-β-lactamases (SBLs and MBLs) produced by multidrug resistant bacteria (3, 4). While combinations of β-lactam and SBL inhibitors are on the market, no approved therapies combined β-lactams with specific MBL inhibitors (5, 6). APC148 is a novel selective zinc-inhibitor targeting Ambler class B MBLs (e.g. NDM, VIM, IMP) that restores β-lactam antibiotic activity without having intrinsic antibacterial effects (7). As both SBLs and MBLs can be expressed in the same pathogen, simultaneous inhibition of both enzymes is essential (8, 9, 10).

In an *in vitro* susceptibility testing of a global collection of 176 MBL and SBL producing Enterobacterales isolates (SENTRY, 2019-2022) APC148 was tested in combination with either cefepime-avibactam or meropenem-avibactam and compared to pipeline and approved antibiotics. APC148 showed superior activity against all other products with a MIC_90_ of 0.12 µg/mL. (11, 12). This high activity of meropenem-avibactam with APC148 in multidrug resistant pathogens was recently confirmed in an Indian collection of 303 isolates of *E. coli* and *K. pneumoniae* (11, Internal report, 2026). These *in vitro* results have been confirmed by *in vivo* studies and support the development of APC148 for the treatment of patients with serious infections caused by multidrug-resistant Gram-negative pathogens.

Based on preclinical data, a first-in-human study was conducted to evaluate the safety, tolerability and pharmacokinetics of single ascending doses of APC148 in a randomized, placebo-controlled study in healthy adults.

## RESULTS

### Disposition and demographics

A total of 46 participants were randomized into six escalating dose cohorts. Cohort 1 included six participants, four randomized to APC148 and two to placebo. Cohorts 2 through 6 each included eight participants, six randomized to APC148 and two to placebo. All participants received a single 3 h intravenous infusion of APC148 or placebo and completed the study according to protocol. Demographic characteristics are summarized in Table

1. Baseline characteristics were generally comparable across the cohorts. Overall, a higher proportion of men than women was enrolled.

### Safety and tolerability

All participants tolerated APC148 well. All adverse events were of mild intensity. There were no deaths, serious adverse events, or adverse events leading to study- or treatment discontinuation. There were no clinically relevant mean changes from baseline in safety laboratory parameters, vital signs or physical examinations between APC148 and placebo, and no apparent trends indicating dose-dependent relationship with respect to these safety parameters.

Except for one instance of erythema, all local tolerability adverse events were assessed as being associated with the study procedures and not APC148.

A total of five adverse events, all of mild intensity, were assessed as related to APC148, including erythema, rash, muscular weakness, headache and a transient and asymptomatic electrocardiogram (ECG) QT prolongation. The ECG QT prolongation measured using heart rate corrected QT interval (QTcF) was observed in one female participant in the 760 mg dose cohort. (Table 2). The maximum QTcF value was 469 ms, observed 3 h and 30 min after start of infusion. On the day prior to infusion, QTcF was 446 ms, and at pre-dose, it was 433 ms.

**Table 1.** Demographics.

| Parameter | Placebo<br>(n = 12) | APC148 dose |  |  |  |  |  |
| --- | --- | --- | --- | --- | --- | --- | --- |
|  |  | 50 mg<br>(n = 4) | 150 mg<br>(n = 6) | 300 mg<br>(n = 6) | 450 mg<br>(n = 6) | 650 mg<br>(n = 6) | 760 mg<sup>1</sup><br>(n = 6) |
| Sex [n (%)] |  |  |  |  |  |  |  |
| Female | 4 (33) | 2 (50) | 2 (33) | 0 | 1 (17) | 2 (33) | 1 (17) |
| Male | 8 (67) | 2 (50) | 4 (67) | 6 (100) | 5 (83) | 4 (67) | 5 (83) |
| Age (yrs) [mean (SD)] | 39.1 (14.4) | 52.3 (9.6) | 50.7 (9.4) | 31.0 (9.1) | 39.0 (15.1) | 40.0 (15.3) | 34.7 (13.6) |
| Race [n (%)] |  |  |  |  |  |  |  |
| White | 12 (100) | 4 (100) | 4 (67) | 6 (100) | 6 (100) | 6 (100) | 5 (83) |
| Asian |  |  | 1 (17) |  |  |  | 1 (17) |
| Multiple |  |  | 1 (17) |  |  |  |  |
| Weight (kg) [mean (SD)] | 75.0 (9.1) | 75.3 (4.7) | 81.1 (20.1) | 82.0 (9.6) | 77.4 (9.4) | 75.9 (10.6) | 73.0 (7.1) |
| BMI (kg/m<sup>2</sup>) [mean (SD)] | 23.9 (3.0) | 24.7 (3.0) | 26.2 (4.1) | 24.6 (3.3) | 25.3 (3.3) | 24.5 (2.4) | 23.5 (3.3) |

1) Following dose group 5 (650 mg), where one participant approached the estimated maximum exposure of APC148, the increment to the next dose group was reduced to 760 mg.

**Table 2.** Subjects with at least one related adverse event.

| Adverse event | No. (%) for each group |  |  |  |  |  |  |  |
| --- | --- | --- | --- | --- | --- | --- | --- | --- |
|  | Placebo<br>(n = 12) | APC148 dose |  |  |  |  |  |  |
|  |  | 50 mg<br>(n = 4) | 150 mg<br>(n = 6) | 300 mg<br>(n = 6) | 450 mg<br>(n = 6) | 650 mg<br>(n = 6) | 760 mg<br>(n = 6) | Pooled<br>(n = 34) |
| Subjects with at least one adverse event |  |  |  |  | 2 (33) | 1 (17) | 2 (33) | 5 (15) |
| Erythema |  |  |  |  | 1 (17) |  |  | 1 (2.9) |
| Rash |  |  |  |  | 1 (17) |  |  | 1 (2.9) |
| ECG QT prolonged |  |  |  |  |  |  | 1 (17) | 1 (2.9) |
| Muscular weakness |  |  |  |  |  | 1 (17) |  | 1 (2.9) |
| Headache |  |  |  |  |  |  | 1 (17) | 1 (2.9) |

### Pharmacokinetics

APC148 demonstrated a consistent pharmacokinetic profile across the 50 mg to 760 mg dose range. Plasma concentrations increased in a dose-proportional manner, as reflected by both C_max_ and AUC, with a mean half-life of 2.3 – 3.0 h (Table 3). Peak concentrations were observed at the end of the 3-h infusion for all dose levels (Figure 1). Mean plasma clearance (CL) was low and dose-independent, and the mean volume of distribution at steady state (Vss) was in the low-to-moderate range.

**Table 3.** Pharmacokinetic parameters for APC148.

| Cohort | Dose (mg) | n | C<sub>max</sub> (µg/mL) | AUC<sub>inf</sub> (h*µg/mL) | CL (L/h) | V<sub>ss</sub> (L) | T<sub>1/2</sub> (h) | F<sub>e</sub> (%) | CL<sub>r</sub><sup>1</sup> (L/h) |
| --- | --- | --- | --- | --- | --- | --- | --- | --- | --- |
| 1 | 50 | 4 | 1.10 (13.3) | 4.78 (11.3) | 10.5 (11.3) | 27.5 (16.5) | 2.3 (12.9) | 38.2 (11.1) | 4.0 (21.6) |
| 2 | 150 | 6 | 3.04 (19.9) | 12.9 (20.7) | 11.6 (20.7) | 32.9 (21.8) | 3.0 (12.5) | 45.4 (6.1) | 5.3 (17.2) |
| 3 | 300 | 6 | 6.72 (15.6) | 25.7 (13.7) | 11.7 (13.7) | 25.4 (16.0) | 2.5 (13.2) | 33.4 (39.4) | 3.9 (52.8) |
| 4 | 450 | 6 | 10.9 (13.8) | 40.3 (14.5) | 11.2 (14.5) | 22.1 (21.0) | 2.4 (14.3) | 63.8 (25.4) | 7.1 (37.9) |
| 5 | 650 | 6 | 15.7 (11.7) | 61.4 (16.0) | 10.6 (16.0) | 24.8 (21.2) | 2.8 (36.9) | 67.9 (16.8) | 7.2 (24.4) |
| 6 | 760 | 6 | 17.2 (9.8) | 63.5 (10.5) | 12.0 (10.5) | 25.4 (16.1) | 2.7 (28.7) | 59.8 (6.9) | 7.2 (13.7) |

C<sub>max</sub>, maximum plasma concentration; AUC<sub>inf</sub>, area under the plasma concentration versus time curve from time zero extrapolated to infinity; CL, total body clearance; V<sub>ss</sub>, volume of distribution at steady state; T<sub>1/2</sub>, terminal elimination half-life; F<sub>e</sub>, Fraction of drug excreted unchanged in urine; CL<sub>r</sub>, renal clearance.
Statistics show geometric means (percent geometric coefficient of variation).
1) Different bioanalytical methods were used for cohort 1-3 and 4-6.

**FIGURE 1.**
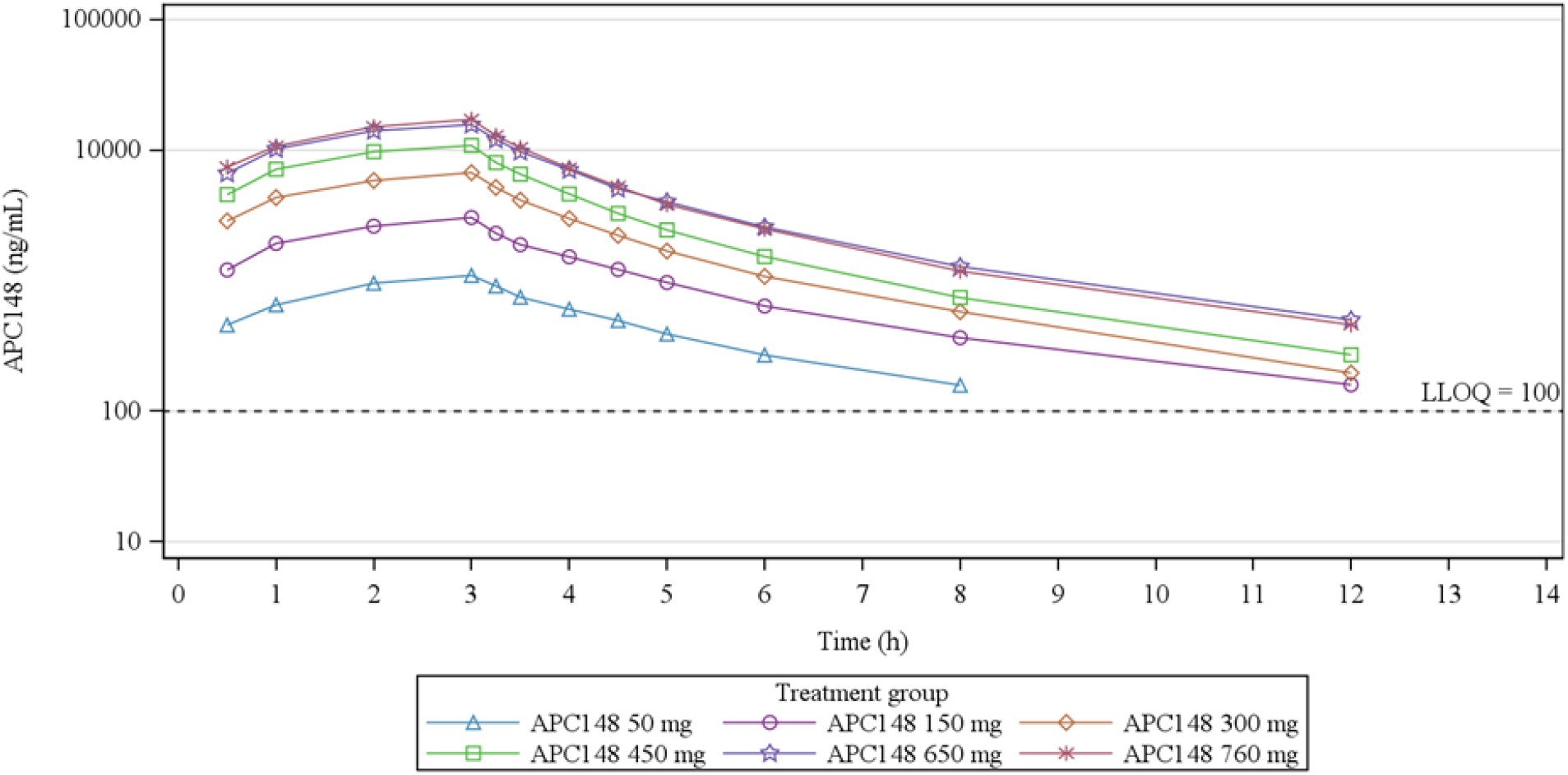
Geometric mean APC148 plasma concentration for cohorts 1 to 6, versus time following a single dose. Logarithmic concentration scale. LLOQ, lower level of detection.

APC148 concentrations were measured in urine fractions collected over 0 – 24 h. The fraction of the administered APC148 dose excreted unchanged in urine (Fe%) ranged from 33% to 68% over the same interval. The mean renal clearance values ranged from 4 L/h to 7 L/h (Table 3).

## DISCUSSION

APC148 is a selective zinc-inhibitor that inhibits MBLs and has no intrinsic antibacterial activity (7). APC148 restores the *in vitro* and *in vivo* activity of meropenem against multidrug resistant MBL producing Enterobacterales (7, Data on file). When meropenem is combined with APC148 and the SBL inhibitor avibactam, the combination shows potent activity against both MBL- and SBL-producing pathogens as shown in *in vitro* and *in vivo* studies (11, 12, Data on file).

As a first step in the clinical development of APC148, a single dose escalating study was conducted in healthy volunteers, to assess safety, tolerability and pharmacokinetics.

Pharmacokinetic analysis indicated an average half-life of 2.6 h, supporting the use of combinations with β-lactam antibiotics such as carbapenems and cephalosporins. The results indicate that renal excretion is the primary route of elimination. This finding is consistent with the animal studies and suggests possible utility in treatment of complicated urinary tract infections. The highest administered dose exceeded the estimated therapeutic dose and was generally well tolerated, with one female in this group presenting a transient, asymptomatic ECG QT prolongation.

These results indicate that APC148 is well tolerated and achieves exposures suitable for combination therapy. The potent inhibition of MBLs observed in preclinical studies, combined with the pharmacokinetics and safety profile, supports further development of APC148.

Combinations of β-lactams antibiotics and β-lactams inhibitors including APC148, addresses a rapidly growing unmet medical need for treatment of patients with MBL-producing infections.

## MATERIALS AND METHODS

### Study design

This was a first in human, randomized, double-blind, placebo-controlled, single ascending dose (SAD) trial of APC148 in healthy adults. The study enrolled participants at a single centre in Sweden between September 2024 and March 2025. The study protocol was approved by the Swedish Medical Products Agency, and the Swedish Ethical Review Authority and was conducted in accordance with the Declaration of Helsinki and Good Clinical Practices. All participants provided written informed consent prior to any study procedure.

A single 3 h intravenous infusion of APCC148 was administered to all participants in 6 sequential cohorts. The administered doses were 50 mg, 150 mg, 300 mg, 450 mg, 650 mg, and 760 mg. Following dose group 5, where one participant approached the estimated maximum exposure of ACP148, the increment of the subsequent dose escalation was reduced.

The first 2 participants in each cohort were dosed in a sentinel fashion, i.e., 1 received APC148 and 1 received placebo as randomized. The safety and pharmacokinetic data for a completed dose cohort were reviewed by a safety review committee before making the decision to escalate to the next dose cohort.

### Participants

Healthy males and females of non-child-bearing potential, aged ≥18 to ≤60 years, with a body mass index ≥ 18.5 and ≤ 30.0 kg/m^2^, and adequate renal function were considered eligible for trial participation. A participant could only be enrolled within 1 dose cohort.

### Safety assessments

Safety was assessed based on the occurrence of adverse events, local tolerability, evaluation of vital signs, 12-lead electrocardiogram (ECG), laboratory parameters (chemistry, haematology, coagulation, urinalysis) and physical examination that was performed throughout the study. Safety was followed for 7 days post-dose.

### Pharmacokinetic assessments

Blood samples for determination of APC148 in plasma were collected pre-dose and at 30 min, 1, 2, 3, 3.25, 3.5, 4, 4.5, 5, 6, 8, 12, 24 and 48 h after the start of drug administration. For all cohorts, urine was collected pre-dose and at intervals of 0 to 6, 6 to 12 and 12 to 24 h after the start of the administration.

### Bioanalytical methods

Human plasma with K2EDTA (dipotassium ethylenediaminetetraacetic acid) were processed using protein precipitation and assayed using the UPLC-MS/MS system. The calibration ranges of 100 to 150,000 ng/ml were used. Urine samples were also analysed using the UPLC-MS/MS method. The bioanalytical method used for the urine analysis of samples from cohort 1-3 (50-300 mg) was fully validated over the concentration range of 100 to 150 000 ng/ml. Due to several urine samples of cohort 4 (450 mg) exceeding the upper limit of quantification a new exploratory method was developed in the concentration range of 1 000 to 2 000 000 ng/mL and applied for samples collected in cohorts 4 to 6 (450-760 mg).

### Pharmacokinetic analysis methods

Individual participant plasma and urine PK parameters were evaluated using non-compartmental analysis methods (Phoenix WinNonlin®, Certara, USA). Calculated plasma PK parameters were among others, C_max_, AUC_0–inf_, CL, V_ss,_ and T_1/2._ Non-compartmental analysis was based on the actual sampling times recorded during the trial. The urine PK parameters were Ae (amount of unchanged drug excreted in urine during the 0 to 24h interval), Fe (fraction of IV administered drug that is excreted into urine) and CL_R_ (renal clearance).

## Data Availability

All data produced in the present study are available upon reasonable request to the authors

## ACKNOWLEDGEMENTS

The study was supported by the Eurostars Programme (Project No. 132), and co-funded by the European Union and national funding authorities. Bioanalysis was performed with Lablytica AB, Uppsala, Sweden. All authors contributed to data analysis and interpretation, as well as manuscript review.

